# Utility of Plasma Cell-free Chromatin Immunoprecipitation to Detect Cardiac Allograft Rejection

**DOI:** 10.1101/2025.03.04.25323391

**Authors:** Moon Kyoo Jang, Oladele Oluwayiose, Neelam Redekar, Temesgen E Andargie, Woojin Park, Muhtadi Alnababteh, Tom Hill, Kellie Phipps, Hyesik Kong, Xin Tian, Helen I Luikart, Michael A Solomon, Palak Shah, Hannah A Valantine, Kiran Khush, Sean Agbor-Enoh

## Abstract

**Background:** Antibody-mediated rejection (AMR) remains the major risk factor for allograft loss across all solid organ transplantation. Unfortunately, its diagnosis relies on biopsy, an invasive gold standard that often sample unaffected allograft tissue leading to missed diagnosis. Plasma donor-derived cell-free DNA (dd-cfDNA) is noninvasive biomarker that has high sensitivity but low specificity for AMR diagnosis. This proof-of-concept study assessed the utility of cell-free chromatin immunoprecipitation (cfChIP) as a surrogate for gene expression to detect cardiac AMR and the associated pathobiology.

**Methods:** The discovery GRAfT multicenter cohort of heart transplant patients (NCT02423070) identified AMR, acute cellular rejection (ACR), and stable controls based on biopsy and dd-cfDNA results. Plasma cfChIP-sequencing was performed to identify peaks, associated genes and pathobiological pathways. Plasma from an external cohort (GTD, NCT01985412) was also analyzed to verify pathways identified. Digital droplet PCR (ddPCR) assays targeting differential regions were constructed to test the diagnostic performance of cfDNA to detect AMR/ACR from stable controls (rejection-specific assays) or AMR from ACR (AMR-specific assays).

**Results:** The cohort included 21 AMR, 28 ACR, and 45 stable controls from GRAfT and GTD, and 23 healthy controls. cfChIP detected expected active genes, including housekeeping genes and gene targets of transplant immunosuppressive drugs but not inactive genes. Unsupervised clustering of the discovery GRAfT cohort assigned 95% of samples correctly as AMR, ACR or stable control. Differential analysis identified pathobiological pathways of AMR such as neutrophil degranulation and complement activation. The pathways were consistent in GTD samples. Rejection-specific assays detected AMR/ACR from controls with AUC of 0.78 - 0.95. AMR-specific assays detected AMR from ACR with AUC of 0.71 - 0.85, sensitivities of 0.73 - 0.94 and specificities of 0.73 - 0.80.

**Conclusion:** This study provides valuable preliminary data supporting the use of cfChIP to detect AMR and the associated pathobiological pathways.

## Introduction

Timely diagnosis and effective treatment of antibody-mediated rejection (AMR) remain important cornerstones of post-heart transplant care (1). The endomyocardial biopsy remains the invasive gold standard to obtain allograft tissue for AMR diagnosis; unfortunately, the biopsy could sample unaffected allograft tissue since it performed without guidance to target affected tissue. The biopsy samples are analyzed by histopathology, which is limited by low sensitivity and high interobserver variability (2). Consequently, the biopsy approach often leads to delayed or missed diagnosis, even in situations when patients present with significant reduction in allograft function. While biopsy-based gene expression profiling and plasma donor-derived cell-free DNA (ddcfDNA) show better sensitivities, the approaches either rely on biopsy to obtain tissue samples or show low specificity for AMR versus acute cellular rejection (ACR), another phenotype of acute rejection that requires different therapeutic approaches.

Cell-free DNA are short DNA fragments released during cell death into the circulation. The first-generation cfDNA assays for rejection monitoring target transplant donor-recipient genetic differences to identify and quantitate plasma donor-derived cfDNA (dd-cfDNA) (3). This non-invasive approach demonstrates low specificity but high negative predictive value for AMR/ACR, making it ideal for surveillance monitoring (4,5). In the dd-cfDNA surveillance strategy, a negative test gives a reassuring “all clear” while a positive test gives an “Alarm” signal requiring further evaluation with biopsy (6). The combined dd-cfDNA/biopsy monitoring approach is still limited by its reliance on obtaining biopsy tissue. Furthermore, the biopsy is negative in over 75% of cases in which the dd-cfDNA levels are elevated (7). Clearly, novel methods to improve AMR diagnosis and to study its pathobiology are needed.

Epigenetic changes, such as chromatin modification, are known surrogates of gene expression and disease pathobiology (8), and are distinct for AMR, ACR and stable (no rejection) controls. In recent studies show cell-free chromatin immunoprecipitation (cfChIP) targeting H3K4Me3, a chromatin change associated with active gene promoters identified peaks that mapped to expected active but not inactive genes (9,10). The identified genes mapped to biologically plausible pathways associated to disease. In this study, we test if cfChIP targeting H3K4Me3 detects the pathobiological pathways of AMR and distinguishes AMR, ACR, and stable controls.

## Methods

### Patient recruitment

The primary analysis performed cfChIP on patient samples obtained from an ongoing multicenter prospective cohort study (GRAfT; ClinicalTrials.gov" NCT02423070). Samples from an external single center cohort study (GTD; ClinicalTrials.gov" NCT01985412) were also analyzed to verify the biologic plausibility of the pathways identified by cfChIP. GRAfT and GTD recruit heart transplant patients to assess the utility of dd-cfDNA to detect acute rejection. Participating centers in both studies use surveillance endomyocardial biopsies and clinically indicated biopsies to monitor patients for AMR and ACR. Patients also have routine echocardiograms to assess allograft function. Patients were maintained on a triple immunosuppression regimen of prednisone, tacrolimus, and mycophenolate, prednisone is often tapered off by 6 – 12 months post-transplant (11). GRAfT and GTD collect plasma samples at time of surveillance biopsies for dd-cfDNA testing (7,26), using similar sample collection protocols (7,26). Healthy controls were included as a comparator group. Healthy controls were recruited at the time of blood donation at the NIH Clinical Center under a Department of Transfusion Medicine protocol (ClinicalTrials.gov" NCT00001846). All patients provided a written informed consent. The study was approved by the institutional review boards of the NIH and participating centers.

### Definition of AMR, ACR and stable controls

Center pathologists reviewed biopsy slides to grade AMR and ACR using the International Society for Heart and Lung Transplant Consensus Guidelines (27,28). Given the low sensitivity and high interobserver variability in biopsy grading, this proof-of-concept study included high grade AMR (≥ pAMR2) or low-grade AMR (pAMR1) plus %dd-cfDNA>0.25% and high grade ACR (grade 3R) or low grade ACR (grade 2R) plus %dd-cfDNA>0.25%. Stable controls included samples with AMR and ACR biopsy grade 0 and no reduction in left ventricular ejection fraction on echocardiography.

### Cell-free chromatin immunoprecipitation and sequencing library construction

Chromatin immunoprecipitation (ChIP) and library preparation for cfDNA were performed with minor modifications to previously published protocols (9,29). Briefly, magnetic beads with conjugated antibodies were incubated with 0.7 ml of plasma and treated with 1X protease inhibitor cocktail (Roche) overnight at 4°C. Beads were magnetized and washed on ice. After elution, histone protein components were digested via proteinase K (Epicenter) treatment. ChIPed cfDNA was purified using a 1.4X SPRIselect (Beckman Coulter) and used to prepare DNA library for NGS sequencing using the Accel-NGS 2S Plus DNA Library Kit (IDT) with unique dual indexing according to the manufacturer’s instructions. Input cfDNA and NGS sequencing protocols were also reported previously (5,7,8).

### Sequencing data analysis

The cfChIP sequencing data was processed using default parameters with the ‘chrom-seek’ workflow (https://github.com/OpenOmics/chrom-seek). Briefly, sequencing reads were trimmed (30) and mapped to ENCODE hg19 v1 (31). Unaligned reads were removed (‘SamToFastq’ function in Picard (https://broadinstitute.github.io/picard/)). Sequencing saturation was predicted using Preseq (v3.1.2) (32). Reads with low quality (mapQ) were filtered out using SAMtools (v1.17) (33). PCR duplicates were removed with ‘MarkDuplicates’ function in Picard (v2.27.3). Read alignments were visualized in BigWig format and the data was normalized by reads per genomic content (RPGC) using deepTools (v3.5.1) (34). Background signal was removed by subtracting the normalized mapping intensity data of input libraries from those of ChIP libraries across the genome using deepTools with --binSize 25.

Prior to peak calling, reads aligning to chromosomes X, Y, and M were removed using SAMtools. Significantly enriched peaks relative to background were identified with macsNarrow (v2.2.7.1) (12,13) across the cohort for dimensional reduction analyses (described below) (13). This step resulted in selection of all peaks present in at least two libraries within the cohort. Normalized peak counts were log (10)-transformed and used to cluster samples for visualization on Uniform Manifold Approximation and Projection (UMAP). For differential binding analyses, however, we included consensus peaks for at least the number of samples in smaller group in every contrast. All peak counts were normalized using the trimmed mean of *M*-values (TMM) method. Peaks were annotated using Gencode Release 19 (GRCh37) with UROPA (v4.0.2) (14). Differential binding analysis was performed using EdgeR to identify differentially bound peaks significant at FDR<0.05 with at least fold change of 2. Over-representation analysis (p<0.01) of genes located within 10kb window of differentially bound peaks against Reactome pathway database was conducted using clusterProfiler (35) and visualized as a dot plot, with an appropriate background set. Partition distribution was estimated using GenomicDistribution package (v 1.12.0) (36). Data visualizations were generated by the R software (v4.01 or later). Protein-protein interaction (PPI) network was extracted for the genes associated with the enriched pathways using Metascape (37), where PPI were sourced from String (physical score > 0.132) and BioGRID databases. The network was organized and visualized in Cytoscape (v 3.10.2) (38).

### Digital PCR assay (primer-probe) design and testing

Approximately 160 bp chromosomal windows coincident to cfChIP-seq peaks with >5d fold change between AMR, ACR and stable controls were considered for design of ddPCR assays (Copy Number Determination tool in the Assay Design Engine, Bio-Rad). The designed and synthesized ddPCR assays were tested for performance using cfChIP DNA libraries and input cfDNA libraries. Certain amount (∼0.354 ng) of DNA libraries were used in the ddPCR assay with a set of Diff ddPCR assays, labeled with FAM or HEX. cfChIP DNA and input cfDNA were measured separately, and the fold enrichment was calculated by dividing the copies of cfChIP DNA by the copies of input cfDNA. The fold enrichment of the experimental differential target was further adjusted by dividing it by the fold enrichment of positive controls. Digital PCR assays were performed for AMR, ACR, and no rejection controls.

### Statistical Analysis

Statistical analysis was performed using GraphPad Prism 9.3.1 and R for Statistical Computing (2023). Data are presented as medians with interquartile ranges (IQR) for continuous variables. Mann-Whitney U test was used to compare two groups with correction for multiple comparison (Dunn’s multiple comparison test). The area under-the-receiver-operator characteristics curve (AUC) was computed using *C-*statistics to estimate the diagnostic performance of ddPCR assays.

## Results

### Patient population and rejection events

**Figure 1a** shows the CONSORT diagram and study design. We utilized cfChIP as a surrogate of gene expression to test if plasma cfChIP could detect rejection, identified the two phenotypes and associated pathobiology. The primary analysis included AMR, ACR and stable transplant control samples from the multicenter GRAfT study (GRAfT; ClinicalTrials.gov" NCT02423070). Samples from an external single center cohort study (GTD; ClinicalTrials.gov" NCT01985412) were also analyzed to verify the biologic plausibility of the pathways identified by cfChIP. GRAfT and GTD use surveillance endomyocardial biopsies and clinically indicated biopsies to monitor patients for AMR and ACR. Patients were maintained on a triple immunosuppression regimen of prednisone, tacrolimus, and mycophenolate, prednisone is often tapered off by 6 – 12 months post-transplant (11). Healthy controls were included as a comparator group. Healthy controls were recruited at the time of blood donation at the NIH Clinical Center under a Department of Transfusion Medicine protocol (ClinicalTrials.gov" NCT00001846). All patients provided a written informed consent. The study was approved by the institutional review boards of the NIH and participating centers.

**Figure 1.**
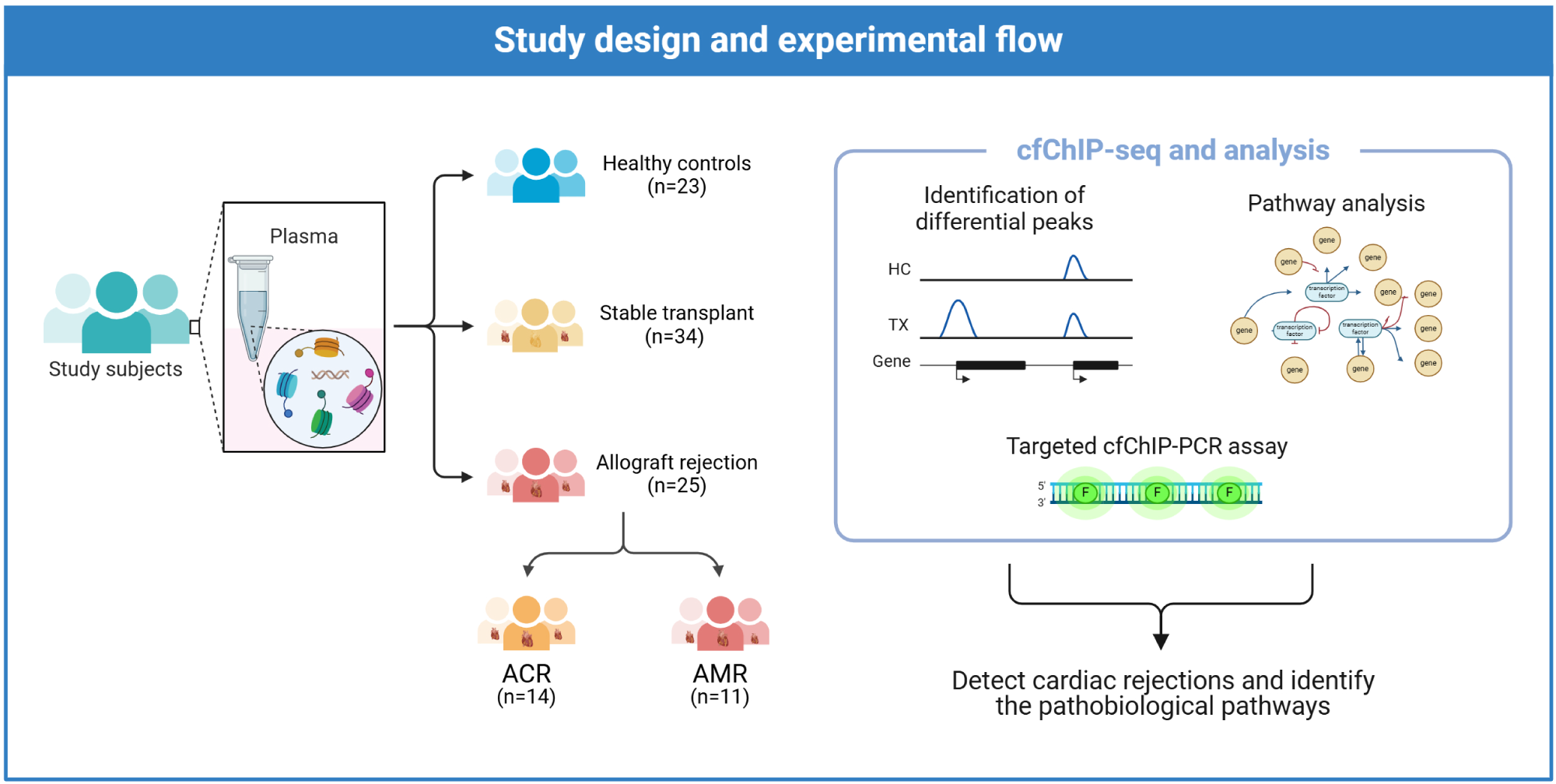
This case-control study of heart transplant patients included AMR, ACR and stable transplant controls. Plasma samples were subject to cfChIP-seq to identify enrichment peaks and associated genes and biological pathways. Primer/probe sets (ddPCR assays) targeting differential cfChIP-seq peaks were constructed to test the performance cfChIP to detect AMR pathways involved in organ dysfunction following heart transplantation.

Given the limitations of biopsy, we select AMR and ACR samples for the GRAfT cohort using biopsy results and %ddcfDNA data (7). We only included AMR samples that showed high grade AMR (≥ pAMR2) on biopsy or low-grade AMR (pAMR1) plus %dd-cfDNA>0.25%, a threshold with high sensitivity to detect AMR/ACR. We selected ACR samples with high grade ACR (grade 3R) on biopsy or low grade ACR (grade 2R) plus %dd-cfDNA>0.25%. After excluding patient with no available plasma samples, recurrent AMR/ACR events within 1 year, or low biopsy-grade AMR/ACR with %ddcfDNA<0.25%, 11 AMR episodes, 14 ACR episodes, and 34 stable control episodes were analyzed for cfChIP-seq from the GRAfT cohort. Median time from transplant to sample collection for the stable controls, ACR and AMR episodes were 194 (17–766), 81 (16–457) and 114 (21–622) days, respectively.

The GTD data had missing %ddcfDNA data. As such AMR/ACR and stable transplant controls were defined using biopsy alone and included AMR grade 1 or higher for AMR and ACR grade 2 or higher for ACR. The study analyzed 10 AMR, 14 ACR, and 11 control samples analyzed from the GTD cohort. Clinical characteristics of patients in the GRAfT and GTD cohorts are shown in **Table 1**. Healthy controls (n =23) were included as a comparator group.

**Table 1:** Demographic and clinical characteristics of patients from the GRAFT and GTD cohorts, and healthy (non-transplant) controls.

| Characteristics | GRAfT cohort (n = 59) |  |  | GTD cohort (n =35) |  |  | Healthy controls<br>(n =23) |
| --- | --- | --- | --- | --- | --- | --- | --- |
|  | Control (n=34) | ACR (n=14) | AMR (n=11) | Control (n=11) | ACR (n=14) | AMR (n=10) |  |
|  | Median(range)/n(%) | Median(range)/n(%) | Median(range)/n(%) | Median(range)/n(%) | Median(range)/n(%) | Median(range)/n(%) |  |
| **Age (years)** | 60 (42 - 70) | 45 (27 - 63) | 51 (27 - 66) | 53 (41 - 69) | 50 (18- 67) | 58 (37 - 60) | 57 (24 – 81) |
| **Sex** |  |  |  |  |  |  |  |
| Male | 30 (88.2) | 8 (57.1) | 9 (81.8) | 10 (90.9) | 10 (71.4) | 5 (50.0) | 9 (39.1) |
| Female | 4 (11.8) | 6 (42.9) | 2 (19.2) | 1 (9.1) | 4 (28.6) | 5 (50.0) | 14 (60.9) |
| **Race** |  |  |  |  |  |  |  |
| White | 24 (70.2) | 8 (57.1) | 2 (19.2) | 3 (27.2) | 9 (64.3) | 5 (50.0) | 17 (73.9) |
| Black | 4 (11.8) | 6 (42.9) | 9 (81.8) | - | 2 (14.3) | 2 (20.0) | 2 (8.7) |
| Others | 4 (11.8) | - | - | 8 (72.9) | 3 (21.4) | 3 (30.0) | 4 (17.4) |
| **Days post-transplant** | 194 (17 - 766) | 81 (16 - 457) | 114 (21 - 622) | 79 (27 - 370) | 138 (27 - 667) | 929 (475 - 1995) | - |

### cfChIP detects gene expression signals in healthy controls and stable heart transplant recipients

Plasma samples for stable transplant controls (SC) of the GRAfT cohort and healthy controls (HC) were analyzed by cfChIP-sequencing to identify consensus peaks. Detailed analysis to identify cfChIP signals, associated genes and differential gene analysis have been reported and shown in **Table S1**. In summary, significantly enriched cfChIP peaks relative to background (12,13) were normalized and used to cluster samples for visualization using the unsupervised Uniform Manifold Approximation and Projection (UMAP). UMAP of consensus peaks (n = 48,800) correctly clustered 100% of healthy controls (HC, n= 23) versus stable transplant controls without rejection (stable controls, SC, n = 33) in the GRAfT cohort (**Figure 2A**). To assessed if cfChIP signals could be interpreted as surrogates of gene expression, we annotated peaks to nearest genes that were within a 10 kb window (14). In both HC and SC, known active genes showed expected H3K4Me3 binding with peaks predominantly localized at the promoter regions of the gene body, which is around the transcription start site. Inactive genes showed no significant binding peaks (**Figure 2B**). The magnitude of peaks correlated with known levels of gene expression. For example, housekeeping genes such as GAPDH that are expressed in all cells showed higher peak magnitude compared to CSF2, which is only expressed in monocytes (**Figure 2C**). We detected 80% (n=7,184) of constitutionally active housekeeping genes in HC and SC (**Figure 2D**). Of the remaining non-housekeeping genes (4,090) identified, approximately 34% were known genes in monocytes and neutrophils (**Figure 2E**), which are cell types that contribute a majority of cfDNA.

**Figure 2.**
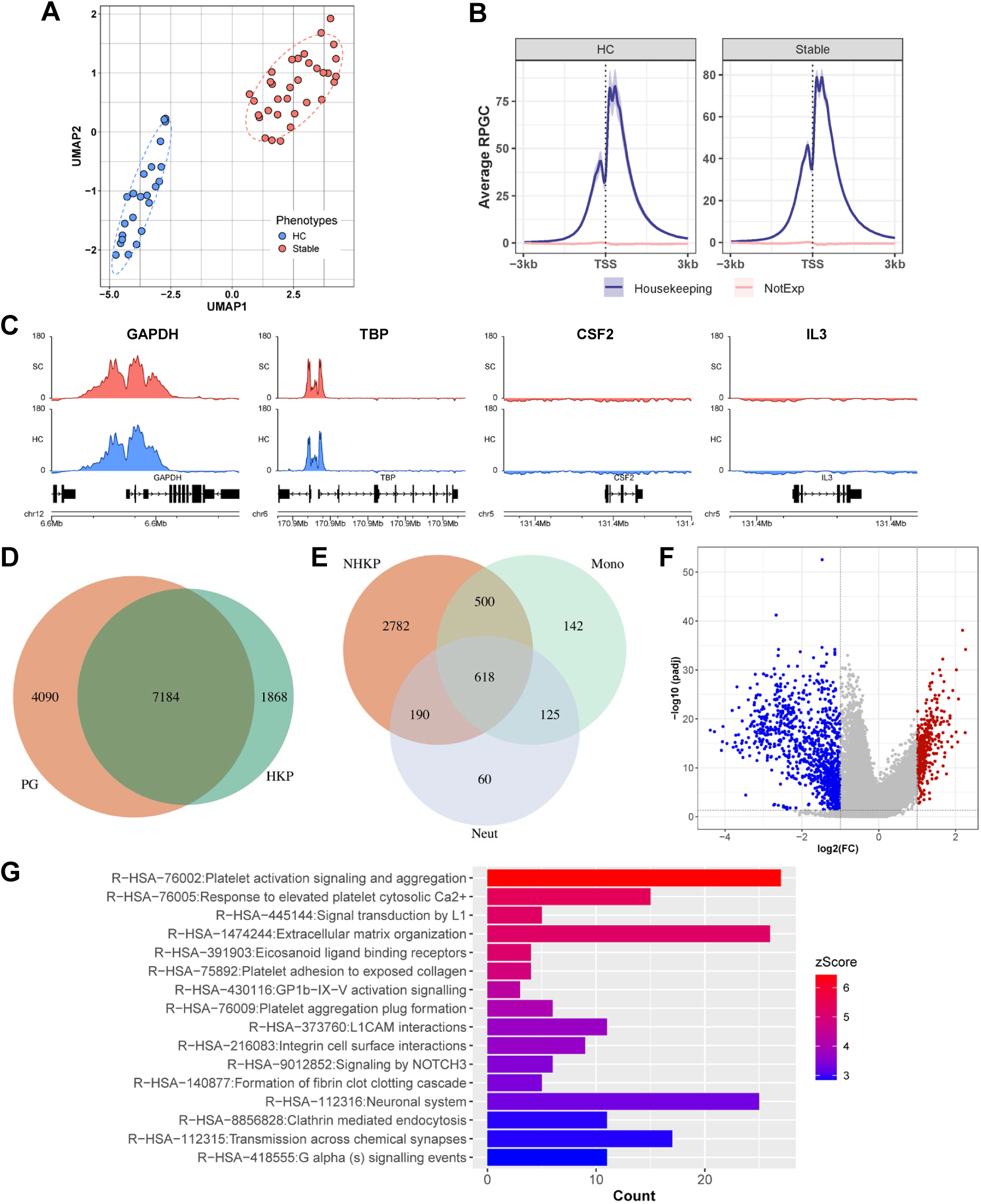
cfChIP profiles of healthy controls and stable transplant patients. (A) UMAP of cfChIP for unsupervised clustering of participants into 2 groups - Healthy (n=23) and stable (n=33) controls. (B) Average coverage plot of cfChIP binding within 3kb up/downstream of TSS for all housekeeping(active) and non-housekeeping (inactive) genes in human genome. (C) Coverage plot of cfChIP binding in few selected active (GAPDH, TBP) and inactive (CSF2, IL3) genes in merged samples. Venn diagram H3K4me3 peaks in stable transplant patients overlapping promoters of constitutively active (D) housekeeping genes and (E) non-housekeeping genes associated to monocytes and neutrophils. (F) Volcano plot of differentially bound peaks (n = 1423) between stable and healthy controls. (G) Bar plot of enrichment analysis to reactome database (p<0.01). Bar length and color represent gene counts and z-score, respectively.

We identify 1,423 differentially bound cfChIP peaks with an FDR<0.05 and at least 2 - fold change between HC and SC (**Figure 2F, Table S1**). A majority of these peaks (n=1,033, 72.6%) displayed decreased binding in SC relative to HC consistent with the immunosuppressed state of SC. Annotation of differentially bound peaks identified 679 unique genes within a 10kb window. Over-representation analyses of these genes in a reactome database showed enrichment of several pathways, including gene targets of transplant immunosuppressive drugs, calcineurin inhibitors (n= 9 genes, CIB1, CHERP, RCAN1, CAMTA1, NR5A1, TBC1D10C, NFATC1, NFATC2, PPP1R1B) and mycophenolate (n = 7 genes, AKT1, PGF, PIK3R5, STK11, TSC1, ULK1, VEGFC), as well as genes related to platelet activation, signaling and aggregation; response to elevated platelet cytosolic Ca2+, formation of fibrin clots (clotting cascade), and extracellular matrix organization (**Figure 2G**).

### cfChIP to detect pathobiological pathways of acute rejection

Plasma samples of AMR, ACR, and stable controls of the GRAfT cohort were analyzed by cfChIP-seq. The unsupervised UMAP of consensus peaks (n=59,512) of GRAfT correctly clustered 95% (n=56/59) of the ACR, AMR and stable control samples analyzed (**Figure 3A**). **Figures 3 B - D** and **Table S2** show differential binding peaks for ACR vs Control (n=3553 peaks, **Figure 3B**), AMR vs Control (n=3603 peaks, **Figure 3C**), and AMR vs ACR (n=596, **Figure 3D**). The genome-wide distributions of the peaks are shown in **Figure 3E**. The differentially bound peaks exhibited gene location that were consistent with known promoters, with approximately 1.5-fold greater enrichment in the promoterCore and 5’ UTR regions, with a minimum of 0.5-fold enrichment in the 3’ UTR and exonic regions and a significant reduction in enrichment at intergenic regions (P < 0.001) (**Figure 3F**). Overall, all differentially bound peaks were enriched within TSS of functional genes (**Figure 3G**).

**Figure 3.**
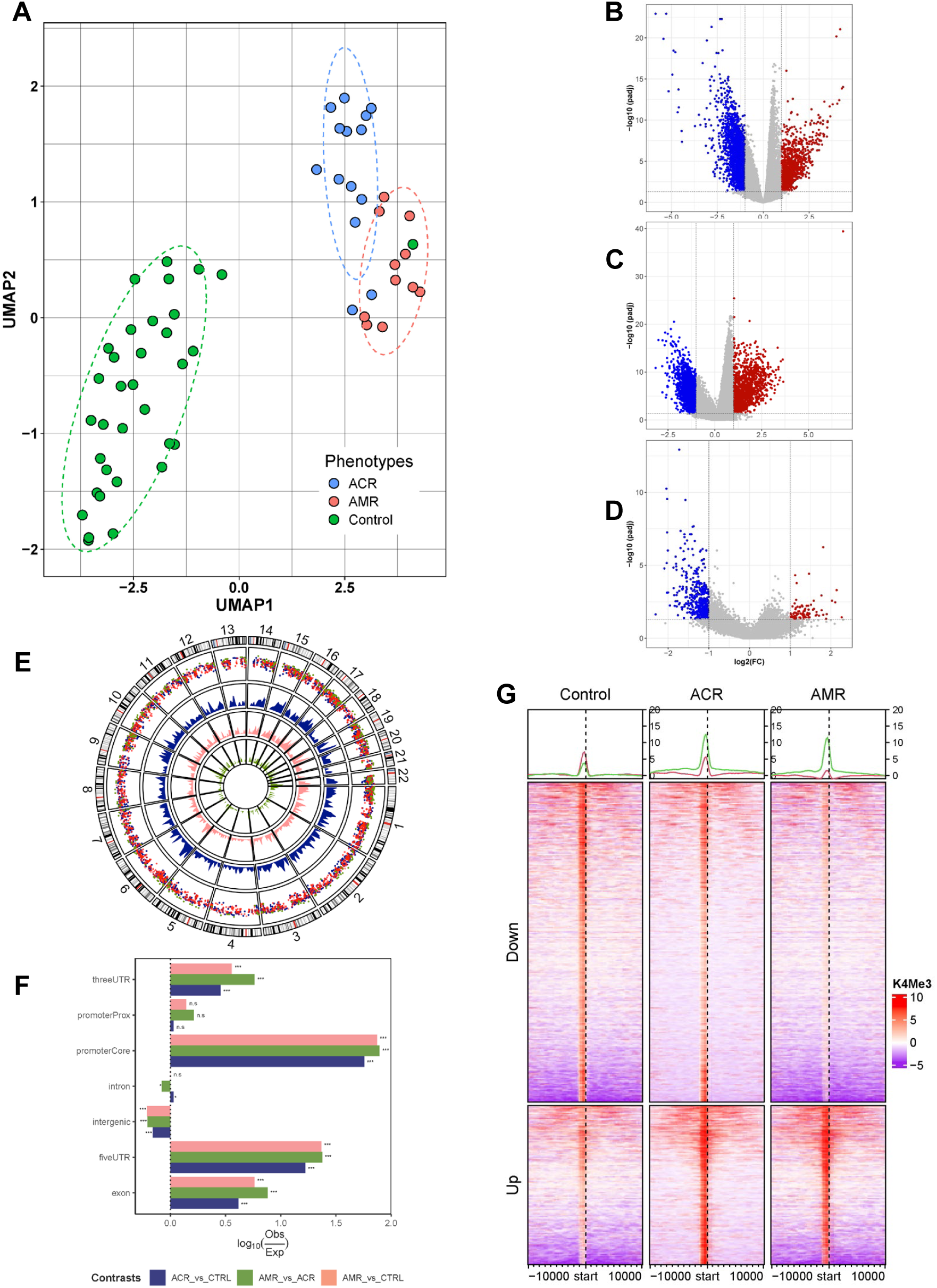
cfChIP profiles of all participating patients in GRAfT cohort. (A) UMAP of cfChIP-seq for unsupervised clustering of patients (n = 59) into 3 phenotypes – Control (n=34), ACR (n=14) & AMR (n=11) based on consensus peaks (n = 59,512, min overlap = 2). (B) Volcano plots of all cfDNA peaks of ACR vs Control, (C) AMR vs Control, and (D) AMR vs ACR. Gray, blue and red datapoints represent non-differentially bound peaks, differentially decreased and increased binding peaks, respectively (FDR<0.05, FC≥2). (E) Circos plot of the distribution of differentially bound peaks (ACR vs Control: 3,553; AMR vs Control: 3,603 ; AMR vs ACR: 596) across chromosomes for all contrasts. (F) Distribution of observed differentially bound peak binding relative to the expected binding across genomic partitions. Fold difference in binding were tested using chi-square where ***p<0.001 & ns p>=0.05. (G) Heatmap of differentially bound peaks around TSS.

The differential cfChIP-seq peaks mapped to 910, 1159, and 221 unique genes within 10 Kb window for ACR vs Control, AMR vs Control, and ACR vs AMR, respectively. Over-representation analyses of these gene sets identified pathways implicated in the pathobiology of AMR and ACR (**Figure 4A, Table S2**). While the AMR vs. controls or ACR vs. controls contrast showed overlapping pathways, the gene targets were different. For example, neutrophil degranulation (R-HAS-6798695) was enriched in all three comparisons. However, the annotated genes were different for ACR vs Control (n= 47) and AMR vs Control (n= 63 genes) with 21 overlapping genes. We also identified pathways related to extracellular matrix organization, calcium mediated platelet activation, GPCR signaling and ligand binding, and lipid/cation flux mediated cardiac conduction – all relevant to the pathobiology of AMR and ACR.

**Figure 4.**
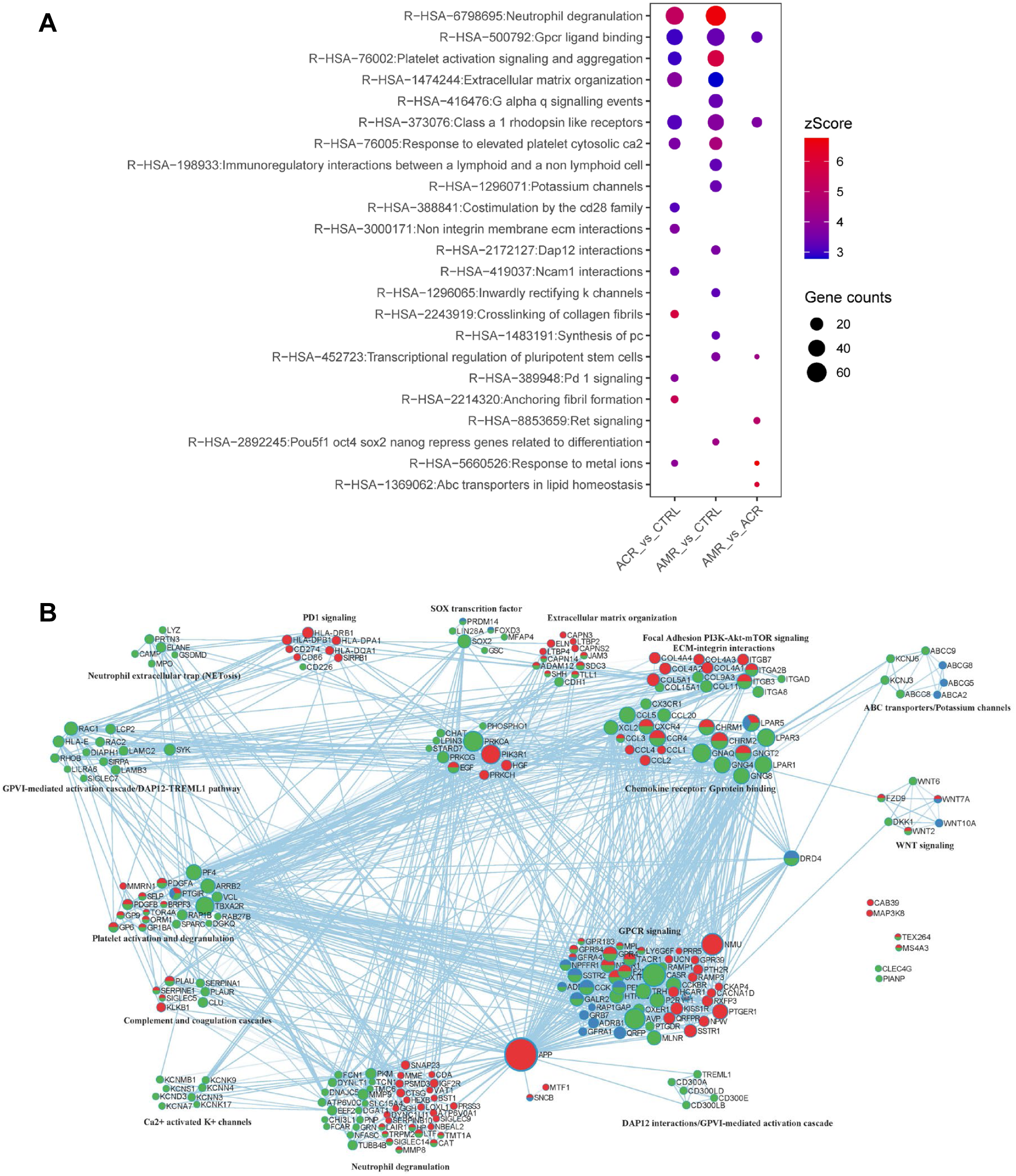
Over-representation analyses of genes located within 10kb of differentially bound peaks across the three contrasts-ACR vs CTRL, AMR vs CTRL and AMR vs ACR. (A) Dotplot of over-representation analysis (ORA) with gene enrichment to reactome (p<0.01) database. Size and color of the dots represent gene counts and zscore respectively. (B) Protein-protein interaction network analysis of genes associated with the ORA.

Protein-protein interaction networks of the differential genes were examined to understand the similarities and differences in pathway inference (**Figure 4B**). Edges (connections between the nodes) in the network are known physical protein-protein interactions from String (physical score > 0.132) and BioGrid (Metascape). Both AMR and ACR groups were associated with genes (or peaks) involved in G-protein coupled receptor (class A1) signaling, neutrophil degranulation, activation of PI3K/Akt-mTOR signaling pathway, extracellular matrix organization and integrin interactions, and platelet activation and aggregation. Despite the occurrence of similar pathways, the physical interactions for the proteins predicted in the ACR and AMR groups were different, as depicted in the network (**Figure 4B**). We also observed classical PD-1 signaling genes associated with the T-cell activation via CD274-MHC II (HLA-DQA1, HLA-DPB1, HLA-DRB1, HLA-DPA1) interaction in ACR group. Unique to AMR group, HLA-binding and Ig-binding receptors on monocytes (HLA-E, LILRA6, CD300LB, CD300LD, CD300E), neutrophil extracellular trap (NET) components (ELANE, MMP9, LYZ, MPO, PRTN3, GSDMD, CAMP), DAP12 interaction components (SIGLEC14, CD300E, CLEC5A, CD300LB, SYK, LCP2, RAC1, HLA-E, TREML1), and SIGLECs (SIGLEC5, SIGLEC7, SIGLEC14) - suggesting immunoregulatory interactions, activation of monocytes/neutrophils and complement cascade associated with severe inflammation and NETosis. In addition to this, we also observed SOX2-mediated transcription of pluripotency in the AMR group, suggesting cell differentiation and proliferation, and activation of potassium channels (KCN and ABC transporters) potentially involved in cardiac conduction. The gene signals identified in AMR vs ACR, on the other hand, were mainly associated with ligand binding and receptors involved in GPCR signaling.

### Concordance of pathways in an independent external cohort

We combined ACR and AMR samples into a unified ‘rejection’ group and compared rejection to stable controls. Given the differences in selection criteria were utilized for AMR. ACR and SC different care practice between centers, we analyzed GRAfT and GTD samples separately. UMAP of identified consensus peaks (n = 59,512 for GRAfT; n = 47,595 for GTD cohort) correctly stratified rejection (AMR+ACR) and stable transplant control groups into separate cohorts for both GRAfT and GTD (**Figure S1 A & B, Table S3**). Differential binding analysis revealed 1,705 and 968 differentially bound peaks between the rejection group and stable controls in the GRAfT and GTD cohorts, respectively, with 170 peaks shared between the two cohorts (**Figure S1C**). While overlapping peaks was a low number, the differentially bound peaks exhibited reduced binding in the rejection group compared to controls for both cohort (GRAfT: 63%; GTD: 66%) (**Figure S1D**). Also, in both cohort, rejection group showed genes enriched in pathways related to neutrophil degranulation and GPCR ligand binding (**Figure S1E**).

Next, we analyzed the GTD samples separately (AMR, n=10, ACR, n=14 and stable controls, n=11) to verify if cfChIP detects biologically plausible AMR or ACR pathways in another cohort. We identified 47,595 consensus peaks, which categorized GTD samples patients into three clinical groups (**Figure S2A**), albeit with more overlap between groups compared to GRAfT (**Figure 3**). Differential binding analysis comparing ACR versus control, AMR versus control, and AMR versus ACR yielded 834, 625, and 16 differentially bound peaks, respectively (FC ≥ 2, FDR < 0.05, **Figure S2B, Table S4**). Overrepresentation analysis using genes associated with these peaks revealed enrichment of pathways similar to those observed in the GRAfT cohort, including neutrophil degranulation, extracellular matrix organization, and GPCR ligand binding (**Figure S2C**).

#### ddPCR assays targeting specific gene loci to detect AMR

The GRAfT and GTD samples were combined to provide a larger cohort size for diagnostic testing. Available plasma samples were subject to cfChIP followed by digital droplet PCR using ddPCR assays that target differentially bound peaks identified on cfChIP-seq (**Table S5**). This included two sets of ddPCR assays, positive and negative controls, to assess the quality of cfChIP. We also designed rejection-specific ddPCR assays that could detect both AMR/ACR relative to SC, as well as AMR-specific and ACR-specific ddPCR assays. Based on estimates from prior analysis, at least 12 episodes of AMR/ACR were needed to sufficiently power the analysis for AMR/ACR versus controls (7).

The combined cohort tested included 47 stable controls, 23 AMR and 21 ACR with available samples. Positive control assays targeted 4 constitutionally active housekeeping genes (ACTB, GAPDH, YWHAZ, SDHA) and showed positive signals in AMR, ACR, and stable transplant control groups (**Figure 5A**). Negative control assays targeted 4 known unexpressed genes (IL3, SMIM42, CSF2, PCP4) and showed negative signals across all groups (**Figure 5B**). Rejection-specific assays target two categories of differential peaks: - 6 assays targeted differential peaks that were increased in stable controls relative to AMR/ACR. These assays detected high signals in stable controls compared to AMR and ACR with an AUC of 0.84 - 0.96 to distinguish controls from AMR/ACR (**Figure 5C**, **Figure S3A**, **Table 2**). There were 6 rejection-specific assays targeting peaks that were increased in AMR/ACR relative to stable controls. These assays detected higher signals in AMR and ACR compared to stable controls and showed AUC of 0.90 - 0.94 to detect AMR/ACR from stable controls (**Figure 5D, Figure S3B, Table 2**). AMR-specific assays targeted differential peaks that were increased for AMR relative to stable controls and ACR. These assays detected higher peaks in AMR compared to ACR and controls. The assays showed AUC of 0.92 - 0.95 to detect AMR from combined ACR/controls (**Figure 5E, Figure S3C, Table 2**). ACR-specific assays generally showed lower peak signals on cfChIP-seq and low ddPCR performance to differentiate ACR with AMR or controls (data not shown).

**Figure 5.**
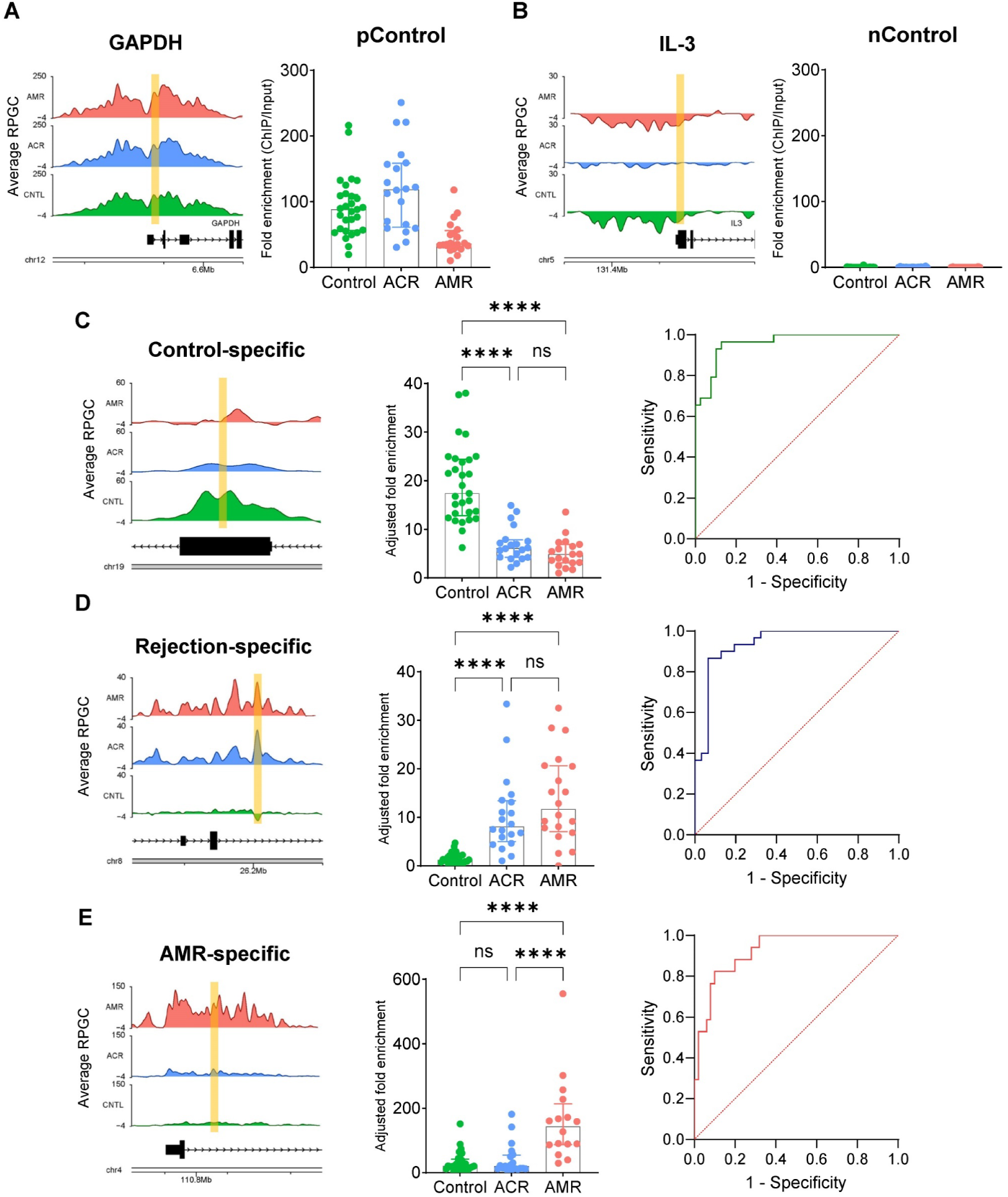
Identification of allograft rejection in heart transplants through the analysis of cfChIPed DNA library using ddPCR. (A) and (B) Average coverage plots of cfChIP binding around the promoter regions of the active gene GAPDH (A) and the inactive gene IL-3 (B), used as positive and negative controls, respectively. The targeted regions for ddPCR assays are marked in yellow (left). cfChIP ddPCR data on the targeted promoter regions of GAPDH and IL-3 for stable control, ACR, and AMR patients. The data were normalized by calculating the ratio of cfChIP DNA signals to input cfDNA signals (right). (C)-(E) Representative coverage plots of cfChIP binding around the control-specific (C), rejection-specific (D), and AMR-specific regions (E). The targeted regions for ddPCR assays are marked in yellow (left). cfChIP-PCR data from cfChIP DNA libraries, enriched from plasma samples of stable control, ACR, and AMR patients, are shown. The data were normalized by calculating the ratio of cfChIP DNA signals to input cfDNA signals, followed by further adjustment by dividing by the average of normalized cfChIP-PCR signals from four positive controls (center). Receiver Operating Characteristic (ROC) curve analysis for predicting stable control (C), allograft rejection (D), and AMR (E) in heart transplant patients based on cfChIP-PCR analysis with specific ddPCR assays (Right).

**Table 2:** Diagnostic performance of ddPCR assays (primer/probe sets)

| Assay ID | Detection_Positive | Detection_Negative | AUC | Threshold | Sensitivity | Specificity | PPV | NPV |
| --- | --- | --- | --- | --- | --- | --- | --- | --- |
| Control1 | CNTL | Rejections | 0.96 | 9.69 | 0.97 | 0.87 | 0.85 | 0.97 |
| Control2 | CNTL | Rejections | 0.89 | 7.08 | 0.83 | 0.85 | 0.81 | 0.87 |
| Control3 | CNTL | Rejections | 0.88 | 8.07 | 0.87 | 0.78 | 0.74 | 0.89 |
| Control4 | CNTL | Rejections | 0.94 | 14.68 | 0.87 | 0.88 | 0.84 | 0.90 |
| Control5 | CNTL | Rejections | 0.95 | 9.05 | 0.97 | 0.88 | 0.85 | 0.97 |
| Control5 | CNTL | Rejections | 0.84 | 8.10 | 0.90 | 0.75 | 0.73 | 0.91 |
| Rejection-1 | Rejections | CNTL | 0.94 | 4.38 | 0.85 | 0.96 | 0.97 | 0.82 |
| Rejection2 | Rejections | CNTL | 0.90 | 11.14 | 0.83 | 0.83 | 0.87 | 0.78 |
| Rejection3 | Rejections | CNTL | 0.94 | 20.04 | 0.90 | 0.87 | 0.90 | 0.87 |
| Rejection4 | Rejections | CNTL | 0.91 | 2.96 | 0.87 | 0.87 | 0.89 | 0.84 |
| Rejection5 | Rejections | CNTL | 0.94 | 8.26 | 0.87 | 0.90 | 0.92 | 0.84 |
| Rejection4 | Rejections | CNTL | 0.91 | 4.39 | 0.80 | 0.93 | 0.94 | 0.78 |
| AMR1 | AMR | CNTL+ACR | 0.92 | 86.52 | 0.81 | 0.90 | 0.72 | 0.94 |
| AMR2 | AMR | CNTL+ACR | 0.94 | 16.13 | 0.90 | 0.94 | 0.86 | 0.96 |
| AMR3 | AMR | CNTL+ACR | 0.92 | 10.04 | 0.95 | 0.82 | 0.67 | 0.98 |
| AMR4 | AMR | CNTL+ACR | 0.95 | 25.38 | 0.95 | 0.90 | 0.79 | 0.98 |

## Discussion

This proof-of-concept study assessed the utility of plasma cell-free chromatin immunoprecipitation (cfChIP) to detect AMR and associated pathobiological pathways, two unmet needs that contribute to poor allograft outcomes. In healthy controls and two heart transplant cohorts, cfChIP identified expected active genes and biological pathways supporting the biological plausibility of cfChIP signals as surrogates for gene expression. The cfChIP detected known pathobiological pathways and pathway interactions for AMR and ACR, further supporting the biologic plausibility. Using sequencing and digital droplet PCR, cfChIP demonstrated excellent diagnostic classification of healthy controls versus stable transplant patients, and of AMR versus ACR versus stable controls. These findings provide crucial preliminary data supporting this approach and suggest the need for further validation studies prior to assessing the clinical utility of cfChIP.

The International Society for Heart and Lung Transplantation (ISHLT) guidelines provide clinical, histopathologic, immunologic, and/or serological assessment criteria for diagnosis of ACR and AMR. ACR is diagnosed as infiltration of recipient mononuclear cells, predominantly T lymphocytes with affinity to IL2, in the myocardial tissue triggering an inflammatory response (15). Activation and proliferation of recipient cytotoxic T-cells within the allograft is a signature of ACR. AMR, however, is diagnosed as infiltration of neutrophils and/or macrophages. There is accumulation of recipient anti-HLA antibodies, fibrin, and complement, in graft capillaries that triggers disruption of vascular endothelial cells (16,17). These hallmark pathways were detected by cfChIP. Involvement of neutrophils in graft rejection is known to start with early infiltration of allograft tissue, to the release of inflammatory cytokines, to activation of the adaptive immune response in both ACR and AMR (18). It is, therefore, not surprising to observe neutrophil degranulation pathways enriched in both ACR and AMR, although the associated genes were strikingly different between the two rejection phenotypes.

Neutrophil granules play a significant role in neutrophil-mediated immunity and their release is tightly regulated by G protein-coupled receptors (GPCR) signaling (19,20). Detection of unique sets of class A rhodopsin GPCR signaling receptors and neutrophil granular contents in the two rejection phenotypes suggest selective regulation of neutrophil-mediated immune responses in ACR and AMR (19). Both the ACR and AMR groups exhibited the classical alloantigen interactions that are expected in T-cell and B-cell mediated immunity, respectively. Detection of antibody-mediated monocyte and neutrophil activation signals (FCAR, CD300, LILRA6), along with TREML1-DAP12 pathway signals (21) may suggest severe pro-inflammatory responses and NETosis in AMR. Detection of components of co-stimulatory (CD86, similar function as CD80) and co-inhibitory (CD274 or PDL1) receptors involved in PD-1 signaling (22) could suggest active regulation of alloreactive T-cell proliferation and survival in ACR. Regulation of PD-1 signaling could shift the transplantation outcome from allograft rejection to tolerance, and several aspects of this signaling are the subject of on-going studies to identify novel immunotherapy targets (23). PDL1 expression in donor tissue has been demonstrated as crucial for allograft tolerance via T-cell inhibition in experimental cardiac transplantation models (24,25). While some of these observations have been reported in biopsies, future studies should assess the mechanistic relevance of these observations.

Our findings support the use of cfChIP to detect AMR and identified key pathobiological pathways associated with AMR and ACR. Sequential use of rejection-specific and AMR-specific ddPCR assays may enable high throughput non-invasive detection and phenotyping of rejection as AMR or ACR. The diagnostic performance of ddPCR assays was consistent with the signal intensity observed on cfChIP-sequencing. Positive control probes targeting constitutively active housekeeping genes, as well as AMR-specific gene loci, showed high signal intensity on cfChIP-seq and high detection performance on ddPCR. The high signal intensity observed with AMR-specific probes is consistent with the pathobiology of AMR as a vascular disease with capillary inflammation and neutrophilic infiltration. Similarly, housekeeping genes are expressed in all tissue types including vascular endothelium and blood cells. As such, disease pathologies with significant intravascular components such as AMR may show higher cfDNA signals compared to diseases that are limited to the organ parenchyma, such as early ACR. This is also consistent with prior studies demonstrating higher levels of dd-cfDNA with AMR than ACR (7). Other factors, such as nuclease digestion within tissues, may contribute to the differences in signal intensity between AMR-specific and ACR-specific primer targets tested. Future studies could better optimize selection of ddPCR assays to improve the diagnostic performance of cfChIP, particularly for ACR.

While our findings support the use of cfChIP to identify pathobiological differences between AMR and ACR with potential diagnostic utility, the study has several limitations. For example, while GRAfT and GTD show overlapping pathobiological pathways, the genes identified showed less overlap. Differences in the sample size between the two cohorts, care practices, rejection grades, and potentially other factors may contribute to lack of greater overlap in the genes identified between GRAfT and GTD. Despite these differences, the concordance observed supports generalizability of our findings even in situations with different care practices. Nonetheless, these findings support the need for future studies to validate the pathobiological pathways identified. These future studies should also compare cfChIP signals between grades of ACR and AMR, as well as patient characteristics or time-dependent factors that could alter cfChIP signals. The future studies could also serve as a diagnostic validation study for cfChIP alone or in combination with other diagnostic approaches. The studies may also define molecular endotypes of AMR, define the effect of current AMR therapies, and the utility of cfChIP to detect treatment response. Pending these future studies, our findings should be interpreted as hypothesis-generating only.

In summary, cfChIP detected biologically plausible surrogates of gene expression, detecting known pathobiological pathways involved in the development of acute rejection after heart transplantation. While AMR and ACR showed overlapping pathways, the gene targets and predictive gene interactions were distinct. If validated in external studies, these findings support the use of cfChIP to non-invasively determine the rejection subtype, which could guide treatment decisions and obviate the need for invasive biopsy procedures.

## Data Availability

The data that support the findings of this study are available from the corresponding author, S.A.E., upon reasonable request.

## Abbreviations

ACR: acute cellular rejection
AMR: antibody-mediated rejection
AUC: area under the curve
ddcfDNA: donor-derived cell-free DNA
ddPCR: Digital droplet PCR
cfChIP: cell-free chromatin immunoprecipitation
ChIP: Chromatin immunoprecipitation
GPCR: G protein-coupled receptors
HC: healthy controls
IQR: interquartile ranges
PPI: Protein-protein interaction
RPGC: reads per genomic content
SC: stable controls
TMM: trimmed mean of M-values
UMAP: Uniform Manifold Approximation and Projection

## Supplemental Material

Figures S1-S3 Tables S1–S5

